# Which functional activities matter most to the patients during the first six months after hip or knee arthroplasty: a survey study

**DOI:** 10.1101/2025.09.26.25336631

**Authors:** Motahareh Karimijashni, Marie Westby, Paul E. Beaulé, Tim Ramsay, Forough Abtahi, Simon Garceau, Stéphane Poitras

## Abstract

**Background:** Understanding what matters most to patients can help clinicians plan healthcare more responsive to patients’ needs after orthopedic surgery. However, there is still little understanding of which activities patients undergoing hip or knee arthroplasty prioritize at different time points after surgery.

**Purpose:** This study aimed to explore key functional activities from patients’ perspectives at different timepoints during the first six months after hip or knee arthroplasty.

**Methods:** A cross-sectional study was conducted at The Ottawa Hospital in Ottawa, Canada. Patients who underwent hip or knee arthroplasty due to osteoarthritis (OA) were recruited. A validated self-report questionnaire based on the activity and participation components of the International Classification of Functioning, Disability, and Health (ICF) core set for osteoarthritis was used.

**Results:** A total of 953 patients participated, including 503 hip arthroplasty patients and 450 knee arthroplasty patients. Patients identified 23 key functional activities and 26 non-key functional activities within 2 weeks, at six, 13, and 26 weeks post-arthroplasty. Patients highlighted different key activities across various stages following hip or knee arthroplasty, with a decline in the number of key activities over time. During the acute recovery phase, patients focused on simpler activities such as walking short distances and self-care, progressing to more advanced activities like squatting and walking long distances as recovery advanced. The findings revealed similar key activities between patients undergoing hip and knee arthroplasty.

**Conclusion:** The study identified how patients’ perspective on key activities changed over different recovery stages following hip or knee arthroplasty. Understanding what matters most to patients enables clinicians to assess outcomes more accurately as well as adapt interventions that align with patients’ needs in order to optimize time to recovery.

## Background

Knee and hip osteoarthritis (OA) are the most common form of joint disease and leading cause of pain and disability.^1,2^ Once conservative treatments fail to control the symptoms, joint arthroplasty is recommended for advanced hip and knee OA.^2–4^ The demand for hip or knee arthroplasty surgeries have increased over the past decades.^5,6^ However, 10% to 30% of patients report little or no improvement in self-reported outcomes, including function and pain after hip or knee arthroplasty.^2–4^ Identifying patients with suboptimal outcomes after hip or knee arthroplasty is challenging due to the lack of consensus on its definition.^2–4^ Ensuring that outcome assessment aligns with patients’ priorities could be key to identifying patients at risk or not optimally recovering and optimal management of outcomes after hip or knee arthroplasty.

Over the past two decades, there has been increasing interest in adopting a more patient-centered approach to assessing treatment outcomes.^8^ Thus, understanding the aspects of care that are important to the patients, and those of lesser importance, is crucial. This knowledge enables healthcare providers to accurately evaluate outcomes and prioritize interventions, thereby enhancing healthcare responsiveness to patients’ preferences and needs. Ultimately, this approach aims to improve patients satisfaction and enhance the overall quality and value of healthcare.^9,10^ Outcome measures need to be informed by patient perspectives to adequately reflect their concerns and needs.^8^ However, previous reviews have indicated that patient involvement in the development process of the tools has been limited.^11,12^

While patients’ expectations and goals before surgery have been extensively investigated in the literature,^14–17^ there is still little understanding of which activities patients undergoing hip or knee arthroplasty prioritize at different time points after surgery. Preoperative information is insufficient for several reasons :1) response shift may occur after arthroplasty, ^18^ as previous studies have shown that patients might revise their functional priorities and expectations and redefine treatment success after hip or knee arthroplasty because their functional status and perception of functional health may change;^19,20^ 2) up to half of patients have overly optimistic attitudes on potential postoperative outcomes which can lead to dissatisfaction after arthroplasty;^17^ and 3) preoperative patient expectations tend to focus only on long-term goals.^19,20^

Patients undergoing arthroplasty due to osteoarthritis (OA) identified improvement in functional outcomes as the main goal of undergoing arthroplasty.^14,15^ Thus, a comprehensive understanding of patient functional priorities at different time points after hip or knee arthroplasty is needed. Therefore, the aim of this study is to determine key functional activities based on patient perspectives within 2 weeks, at 6, 13, and 26 weeks after hip or knee arthroplasty.

## Methods

We conducted a cross-sectional survey and it was reported based on the Strengthening the Reporting of Observational Studies in Epidemiology (STROBE) guideline.^21^ The rationale for choosing this research design is to efficiently reach a large sample in a specific period of time.^22,23^ This study received ethics approval from the Ottawa Health Sciences Network Research Ethics Board (20230252-01H), and the University of Ottawa Board of Ethics (H-06-23-9425).

### Survey questionnaire

In a previous study,^24^ we developed a self-administered questionnaire based on activity and participation components of the International Classification of Functioning, Disability and Health (ICF) core set for OA and established its face and content validity through cognitive think-aloud interviews with 18 patients who underwent hip or knee arthroplasty. This process resulted in 49 close-ended questions on functional activities in the following 12 domains: changing basic body position, maintaining a body position, lifting and carrying objects, walking, moving around, using transportation, driving, washing oneself, toileting, dressing, remunerative employment, and recreation and leisure. We also included an open-ended question to capture any activities that were important to the patient but not listed in our questionnaire.

### Setting and participants

We conducted this study within the orthopedic division at The Ottawa Hospital in Ottawa, Canada, an academic-affiliated institution in an urban setting with a high volume of arthroplasty procedures. The recruitment timing in this study was selected based on recovery pattern after arthroplasty. Within two weeks after hip or knee arthroplasty captures acute recovery. Approximately six weeks postoperative addresses the first post-acute recovery since most adverse events usually occur within 30 days after arthroplasty,^25,26^ and most patients can be considered stable between 4 to 6 weeks after hip or knee arthroplasty. The assessments at 13 and 26 weeks were selected to capture the majority of functional recovery.^27,28^

Participants were adults (18 years or older) who underwent primary elective hip or knee arthroplasty, including total joint arthroplasty, uni-compartmental knee arthroplasty, and hip resurfacing or hemi-arthroplasty due to OA, had surgery one, six, 13, or 26 weeks prior, spoke English, and gave consent to participate in this study. Participants with hearing or speech impairments were included if the research team member (MK) received their consent as it was not required to communicate orally to fill out the questionnaire. Participants who underwent hip or knee arthroplasty for reasons other than primary OA, who had severe cognitive impairment, were unable to read and did not have any support (family member, friend, or caregiver) to read the documents were excluded.

### Sampling and Recruitment

We used in-person and telephone-based recruitment strategies. The patients were recruited during postoperative in-clinic visits with orthopedic surgeons at The Ottawa Hospital or by phone calls. The study used two nonprobability sampling methods, including convenience sampling and purposive sampling. First, patients were recruited using convenience sampling in the first three months of recruitment.^29^ After three months of recruitment, the characteristics of the participants at each time point was compared with the Canadian population (age and gender),^6^ patients at The Ottawa Hospital who underwent hip or knee arthroplasty over the past two years (caregiver support, comorbidities, walking aids), and with findings from relevant research (BMI). This comparison aimed to determine which demographic characteristic was under or overrepresented. In the case of significant differences with the population, purposive sampling was selected for patient recruitment to ensure representativeness. To identify and recruit individuals with underrepresented characteristics, we reviewed patient charts at The Ottawa Hospital. This method allowed us to specifically target and include patients whose characteristics were underrepresented in our sample.

In Canada, all medically necessary procedures, including arthroplasty, are covered under the publicly funded healthcare system. At The Ottawa Hospital, patients receive standardized pre-operative and post-operative care pathways, including rehabilitation services, regardless of socioeconomic status or insurance coverage. As a result, variations in access to care did not influence clinical outcomes and the key post-operative activity measures collected in this study.

### Data collection

After agreeing to participate in this study, the participants had the option to complete the questionnaire either in-person (paper) or online (email). Implied consent was obtained from participants by responding to the survey. For the in-person option, participants were given the choice to receive either a paper version of the questionnaire and consent form physically during their appointments with their orthopedic surgeons or to have them sent by mail. The email contained an embedded link to the survey which directed the interested participants to the implied consent form and survey questions. The Ottawa Hospital REDCap software platform was used to develop the online survey.^30^ In order to increase response rates, non-responders were contacted by a research team member (MK) via telephone approximately 5 and 10 days after receiving the questionnaire.^31^ To identify the non-responders, a research team member (MK) used the participant IDs. The survey could be completed over several sessions and allowed participants to review their answers before the final submission.

In addition, preoperative data including Oxford Knee Score (OKS), Oxford Hip Score (OHS) score, American Society of Anesthesiologists physical status classification (ASA) score and surgical approach were collected from patient charts in Epic and ConEHR. ConEHR is part of the Orthopaedic Surgery Division’s Continuous Quality Improvement program based within The Ottawa Hospital.

### Sample Size

With a margin of error of 5% and a confidence level of 90% based on number of patients underwent hip or knee arthroplasty at The Ottawa Hospital across six months, it was estimated that 123 hip arthroplasty and 112 knee arthroplasty participants for each time period would be representative of the respective patients population at The Ottawa Hospital.^32^ We consider the sample size of patients underwent hip or knee arthroplasty over six months to be aligned with the follow-up period.

### Data analysis

We used the Statistical Package for Social Sciences (IBM SPSS, version 28) to analyze data. Descriptive analyses were used to calculate response rates and participants demographics, such as age, gender, ethnicity, having a caregiver, preoperative function (defined as OKS or OHS^33^), postoperative pain (defined as visual analog scale (VAS) score^34,35^), body mass index (BMI), and comorbidities (defined ASA score^36^). All continuous variables were expressed as means and standard deviations, and categorical variables were reported as frequency and percentage.

With the questionnaire, participants rated each functional activity on four point scales according to its current importance to them and ability to do it.^37,38^ The importance of the activity was dichotomized into: not important (not important, slightly important) and important (important, very important), and ability into: little difficulty (easily, with little difficulty) and a lot of difficulty (with a lot of difficulty, unable to do).

Two research team members (MK and FA) independently coded the open-ended question asking about other important activities with NVivo 12 qualitative data analysis software (QSR International Inc., Burlington, MA, USA).^39^ If a response could align with one of the 49 activities of the questionnaire, it was compared with the corresponding questionnaire response. If there were discrepancies, the coders updated the response accordingly. If a code did not correspond to any of the 49 activities, the coders defined a new code and grouped them to identify uncovered key activities. Two coders met regularly to peer-review the coding process, and discuss interpretations until reaching a consensus.^40^

To identify key functional activities, we considered two factors: the importance of the activity to patients and the difficulty they had performing it. An activity was classified as key if it was both important to patients and challenging for them to carry out. An activity was classified as important if at least 75% of participants rated it as such, consistent with previous studies,^41,42^ and threshold of 15% of participants reporting difficulty with the activity was applied, based on the average proportion of patients experiencing poor functional outcomes after arthroplasty.^43–50^ In addition, we conducted chi-square analyses to compare the importance and difficulty of each activity at each time point between hip and knee arthroplasty. If an activity met the thresholds for hip or knee and there was no statistically significant percentage difference between procedures, we categorized that activity as key for both. A p-value < 0.05 was considered statistically significant.

## Results

From November 2023 to March 2024 and September 2024 to January 2025, 1,170 eligible patients were approached and invited to participate in this study. Of these, 503 patients who underwent hip arthroplasty and 450 knee arthroplasty surgeries completed the questionnaire, resulting in a response rate of 86.6% and 86.5 % for hip and knee arthroplasty respectively. In the last four months of recruitment, we purposely recruited the participants based on age, gender and BMI, aiming to ensure a representative sample of the population. The detailed participant recruitment process is presented in Figure 1.

**Figure 1.**
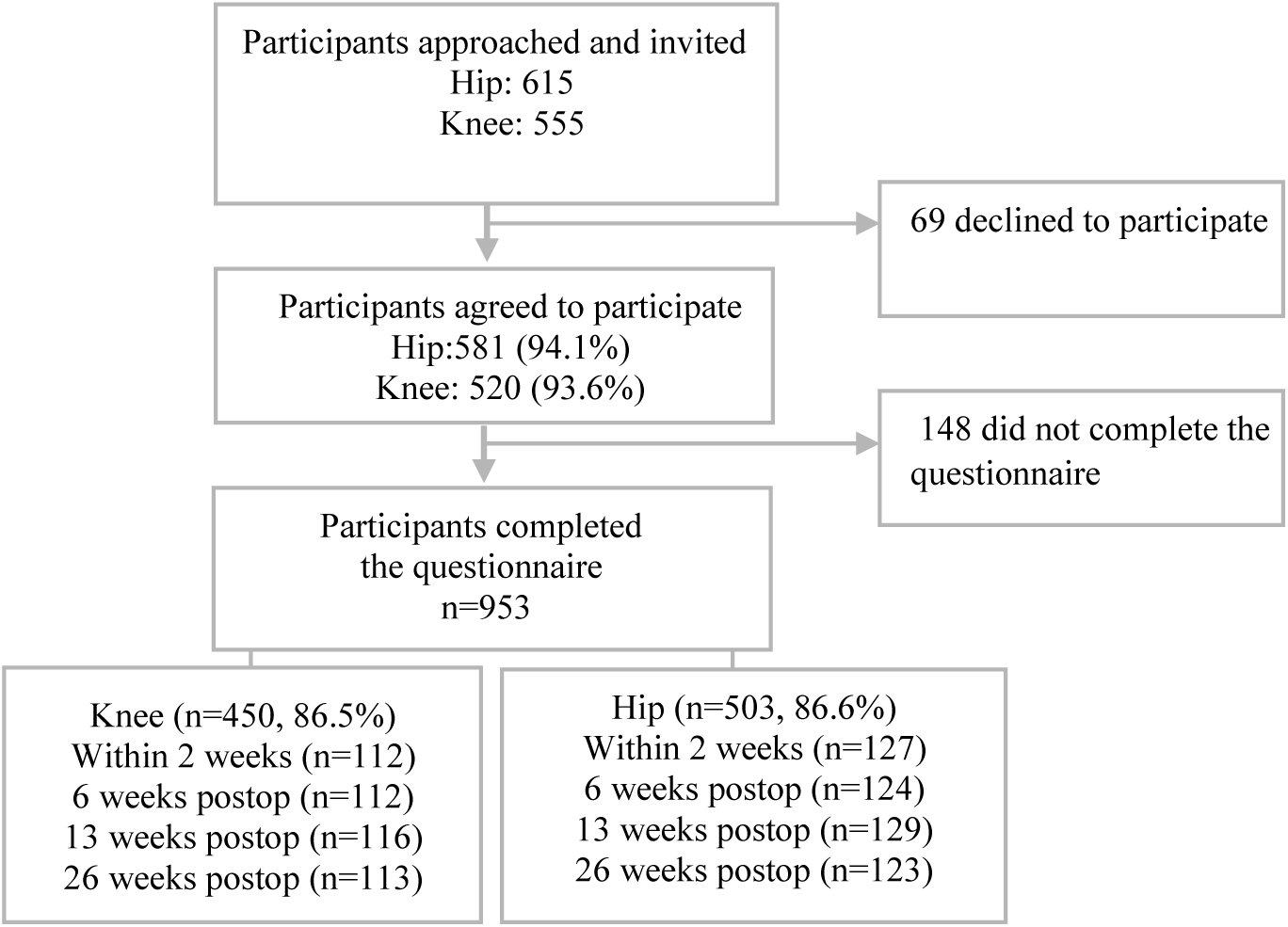
Flowchart of participants recruitment

### Participants Characteristics

Detailed characteristics of the patients at different time points after hip and knee arthroplasty are presented in Appendix A and B, respectively. Patients did not differ significantly across time periods following hip or knee arthroplasty, except in pain levels, use of walking aids after hip or knee arthroplasty, and precautions after hip arthroplasty, which may reflect improvements in recovery over time. When comparing hip and knee arthroplasty, patients who underwent knee arthroplasty were older, had higher BMI, higher preoperative Oxford score, a higher proportion of ASA scores 3 and 4, experienced more pain, and had more contralateral knee joint problems. Patients who underwent hip arthroplasty had more contralateral hip joint problems, with a higher proportion of patients using crutches. Appendix C provides a detailed comparison of the demographic characteristics of patients undergoing hip and knee arthroplasty.

### Key functional activities after hip or knee arthroplasty

Appendix D to G present percentages of importance and difficulty for the 49 functional activities. The results demonstrated different key functional activities across different time points after arthroplasty.

### Key functional activities within 2 weeks after hip or knee arthroplasty

During the first two weeks after hip or knee arthroplasty, 14 key functional activities were identified for both procedures, with the addition of rising from a chair after knee arthroplasty. These activities were related to categories of self-care (n=6), changing and holding a body position (n=5), moving around (n=2), and walking (n=1) (Table 1, appendix D).

**Table 1.** Key functional activities at different time points after hip or knee arthroplasty.

| Activity | 2 weeks postop |  | 6 weeks postop |  | 13 weeks postop |  | 26 weeks postop |  |
| --- | --- | --- | --- | --- | --- | --- | --- | --- |
|  | Knee | Hip | Knee | Hip | Knee | Hip | Knee | Hip |
| **Changing and holding a body position** |  |  |  |  |  |  |  |  |
| Lying down in bed and getting up | ✓ | ✓ |  |  |  |  |  |  |
| Rolling over | ✓ | ✓ |  |  |  |  |  |  |
| Squatting down |  |  | ✓ | ✓ | ✓ | ✓ | ✓ | ✓ |
| Hold a sitting position | ✓ | ✓ | ✓ | ✓ | ✓ | ✓ |  |  |
| Rising from a chair | ✓ |  |  |  |  |  |  |  |
| Hold a standing position | ✓ | ✓ |  |  |  |  |  |  |
| **Lifting and carrying objects** |  |  |  |  |  |  |  |  |
| Lifting a heavy object from the floor |  |  |  |  | ✓ | ✓ | ✓ | ✓ |
| Putting down objects on the floor |  |  | ✓ | ✓ |  |  |  |  |
| Carrying in the arms |  |  | ✓ | ✓ |  |  |  |  |
| **Walking** |  |  |  |  |  |  |  |  |
| Walking short distances | ✓ | ✓ |  |  |  |  |  |  |
| Walking long distances |  |  |  |  | ✓ | ✓ | ✓ | ✓ |
| Walking on different surfaces |  |  | ✓ | ✓ |  |  | ✓ | ✓ |
| **Moving around** |  |  |  |  |  |  |  |  |
| Going up stairs | ✓ | ✓ | ✓ | ✓ |  |  | ✓ |  |
| Going down stairs | ✓ | ✓ | ✓ |  | ✓ |  | ✓ |  |
| **Self-care** |  |  |  |  |  |  |  |  |
| Washing lower body parts | ✓ | ✓ |  | ✓ |  |  |  |  |
| Washing whole body | ✓ | ✓ |  |  |  |  |  |  |
| Drying your body | ✓ | ✓ |  |  |  |  |  |  |
| Putting on clothes | ✓ | ✓ |  |  |  |  |  |  |
| Putting on footwear | ✓ | ✓ | ✓ | ✓ |  |  |  |  |
| Taking off footwear | ✓ | ✓ | ✓ | ✓ |  |  |  |  |
| **Domestic life** |  |  |  |  |  |  |  |  |
| Going shopping |  |  | ✓ | ✓ |  |  |  |  |
| Doing housework |  |  | ✓ | ✓ |  |  |  |  |
| **Community, social and civic life** |  |  |  |  |  |  |  |  |
| Socializing/Participating in community life |  |  | ✓ | ✓ |  |  |  |  |
| Frequency of priorities at each time point | 14 | 13 | 12 | 12 | 5 | 4 | 6 | 4 |

### Key functional activities at 6 weeks after hip or knee arthroplasty

Patients identified 11 key functional activities six weeks after hip or knee arthroplasty, with the additional activity of going down stairs after knee arthroplasty and washing lower body parts after hip arthroplasty. These activities encompassed categories of changing and self-care (n=3), changing and holding a body position (n=4), lifting and carrying objects (n=2), moving around (n=2), domestic life (n=2), walking (n=1), community, social and civic life (n=1) (Table 1, appendix E).

### Key functional activities at 13 weeks after hip or knee arthroplasty

Patients identified four key functional activities 13 weeks after hip or knee arthroplasty, with the additional activity of going down stairs after knee arthroplasty. The activities were related to categories of changing and holding a body position (n=2), walking (n=1), lifting and carrying objects (n=1), and moving around (n=1) (Table 1, appendix F).

### Key functional activities at 26 weeks after hip or knee arthroplasty

Patients identified four key functional activities at 26 weeks after hip or knee arthroplasty. These activities were related to categories of walking (n=2), changing and holding a body position (n=1) and lifting and carrying objects (n=1). Furthermore, two additional activities (going up stairs, going down stairs) were identified after knee arthroplasty (Table 1, appendix G).

### Differences in key activities across the four-time points

Key activities varied across four different time points, with some activities common across various phases. Within the first two weeks after hip or knee arthroplasty, patients identified seven key activities that were not key at later time points, including lying down in bed and getting up, rolling over, rising from a chair, holding a standing position, walking short distances, washing the whole body, drying the body, and putting on clothes. Six key activities identified within the first two weeks after surgery remained key in later stages of recovery, including holding a sitting position, going up stairs, going down stairs, washing lower body parts, putting on footwear, and taking off footwear. As recovery progressed, nine activities that were not identified as key within the first two weeks postoperatively were key six weeks and beyond, including squatting, putting down objects, carrying in the arms, walking on different surfaces, going shopping, doing housework, and socializing. Conversely, holding a sitting position, and putting on and taking off footwear were key at 2, 6 and 13 weeks but not at 26 weeks. Eight activities were key only six weeks after surgery, including putting down objects, carrying in the arms, going shopping, doing housework and socializing. Finally, lifting heavy objects from the floor and walking long distances were key only at 13 and 26 weeks after surgery.

### Other functional activities from qualitative analysis

After qualitative analysis of the open-ended questions from 307 participants, we identified 85 responses that did not match the 49 activities listed in our questionnaire based on the ICF OA core set. Six new ICF activities were identified, including taking care of animals, caring for household objects (vehicle maintenance), other specified moving around (getting into and out of boats or pool), caring for hair, caring for toenails and writing. However, none of them were identified as key activities since they did not meet the thresholds. Table 2 provides the reporting frequency of these activities, shown in aggregate rather than at individual time points.

**Table 2.** Activities identified through qualitative analysis of open-ended questions.

|  | Frequency |
| --- | --- |
| d6506: taking care of animals | 32 |
| d4558: other specified moving around | 10 |
| d650: caring for household objects | 11 |
| d5202: caring for hair | 2 |
| d5204: caring for toenails | 1 |
| d170: writing | 1 |

### Non-key functional activities

The findings indicated that 26 out of 49 activities were not key at any time after hip or knee arthroplasty. Table 3 lists these activities.

**Table 3.** Non-key activities after hip or knee arthroplasty.

|  |
| --- |
| Non-priority activities |
| **Changing and holding a body position** |
| Hold a lying position during the day |
| Hold a squatting position |
| Kneeling down on the ground |
| Hold a kneeling position |
| Sitting on a chair |
| Bending over or to the side |
| Shifting the body from side to side |
| **Lifting and carrying objects** |
| Lifting a light object from the floor |
| Carrying in the hands |
| Carrying on shoulders or back |
| **Walking** |
| Walking around obstacles |
| **Moving around** |
| Going up a curb or step |
| Jumping |
| **Moving around using transportation** |
| Getting in and out of a car |
| Using public transportation |
| Biking |
| Driving |
| **Self-care** |
| Going to the toilet |
| Taking off clothes |
| **Domestic life and Intimate relationships** |
| Assisting others |
| Sexual relationships |
| **Employment** |
| Paid job |
| Unpaid work |
| **Community, social and civic life** |
| Sports |
| Running/jogging |
| Hobbies/ play /Crafts/ Arts and culture |

## Discussion

This study identified key functional activities within the first two weeks, at six, 13 and 26 weeks after hip and knee arthroplasty. Our findings can enable healthcare professionals to tailor care plans to focus on what matters most to the patient at various stages of recovery, promoting patient-centered care.^51,52^ This approach can enhance the effectiveness of postoperative interventions, improve patient adherence to treatment, and ultimately optimize patient outcomes while reducing treatment burden. Patient-centered care is important for individuals experiencing poorer outcomes after hip or knee arthroplasty, where evidence for postoperative intervention is uncertain.^53,54^

To ensure that interventions align with patient key activities, it is essential to incorporate patient-centered outcome measures. Our results can guide the selection of patient-centered outcome measures to evaluate how well patient needs are addressed. Not taking into account patient needs may lead to clinicians missing important treatment outcomes.^55^ Moreover, this study indicated that key functional activities vary over time, with limited differences between hip and knee arthroplasty. This information about patients’ evolving experiences during recovery enable clinicians to manage patient expectations by providing realistic insights into recovery, potentially increasing satisfaction.^56^

The findings highlighted that patients identify different key activities at various stages following hip or knee arthroplasty. When improvements were reached in some key activities, new needs in other key activities appeared. Patients identified simpler key activities during the acute phase of recovery and shifted towards more advanced activities as they progressed. Initially, within the first 2 weeks after hip or knee arthroplasty, patients focused on basic mobility tasks (e.g. rolling over, holding a sitting position, walking short distances), self-care tasks, and dressing. By 6 weeks, there was the shift towards activities requiring increased stability and mobility, such as walking on different surfaces and doing housework, aimed at regaining independence in daily activities. By 13 weeks, key activities advanced to more complex tasks such as walking long distances. Finally, at 26 weeks, key activities consolidated around more demanding tasks, such as squatting down, and lifting a heavy object from the floor. In addition, patients identified some activities as important, but challenging throughout the postoperative recovery such as holding a sitting position, going up and down stairs, squatting, and walking on different surfaces. There were, however, slight inconsistencies, as walking on different surfaces and going up stairs were not key activities at 13 weeks while they were key at six and 26 weeks. These inconsistencies might be due to the margin of error since the results were very close to the cut-offs. Postoperative rehabilitation should prioritize these key activities throughout the recovery to improve performance. However, previous research indicates going up and down stairs is infrequently incorporated into rehabilitation interventions,^57^ and there is limited research available to determine the extent to which other activities are addressed in rehabilitation programs after hip or knee arthroplasty.

Our findings also reveal a decline in number of patients’ key activities over time, indicating a reduction in the important functional activities they had difficulty performing. This suggests that arthroplasty procedures can restore a significant degree of function and enhance independence in the first three to six months after surgery, as demonstrated by previous research.^28,58–60^ However, patients identified six activities after knee arthroplasty and four after hip arthroplasty that were important to them but continued to experience difficulty at 26 weeks after arthroplasty. This is consistent with previous research reporting that 10-30% of patients experience persistent poorer function after arthroplasty,^61^ with a higher proportion of patients reporting poorer functional recovery after knee arthroplasty compared to hip arthroplasty.^48,49^ As previously noted, activities such as squatting, walking on different surfaces, and going up and down stairs pose challenges at all stages of recovery. These findings highlight the importance of implementing targeted rehabilitation strategies early in the recovery process to mitigate these challenges as patients progress.

There was notable overlap in activities between knee and hip arthroplasty which aligns with previous research.^62^ Although this might suggest that similar postoperative rehabilitation interventions could be effective for both hip and knee arthroplasty patients due to similarity in key activities, the treatment strategies to address impairments in these key activities may differ. For instance, while knee range of motion might limit going down stairs in patients who have undergone knee arthroplasty, muscle strength may be the limiting factor for those who have had hip arthroplasty. Our findings did reveal slight differences in key functional activities between patients undergoing hip and knee arthroplasty.^63,64^ Patients who underwent knee arthroplasty reported slightly greater difficulty in performing some key activities. This likely reflects poorer outcomes, as previous studies have shown that patients undergoing knee arthroplasty tend to experience worse outcomes compared to those undergoing hip arthroplasty.^49,65^

Going down stairs at six, 13 and 26 weeks and going up stairs at 26 weeks were key after knee arthroplasty, with a trend towards the cut-off for going up stairs at 13 weeks after knee surgery. These results are consistent with previous research showing that patients who underwent knee arthroplasty had less satisfaction in going up and down stairs compared to those who underwent hip arthroplasty.^66^ This difficulty after knee arthroplasty may be attributed to the higher biomechanical functional demands placed on the knee joints and muscles during stair activities ^67^ and higher ground reaction forces on knees joint.^68^ Additionally, rising from a chair was identified as a key activity within the first two weeks following knee arthroplasty but not after hip arthroplasty. This difference between hip and knee arthroplasty may stem from several factors, such as limited range of motion and quadriceps weakness in patients undergoing knee arthroplasty, which can significantly impact their ability to perform this activity.^69^ Furthermore, washing lower body parts was identified as a key activity at six weeks following hip arthroplasty but not after knee arthroplasty. This might be because washing lower body parts requires a greater range of motion in the hip compared to the knee.^70^

Patients identified 26 ICF OA core set activities that were not key following hip or knee arthroplasty. The findings of key and non-key activities highlight the need to evaluate the content validity of outcome measures used after these procedures because several outcome measures could miss key activities and include non-key activities.^71^ Incorporating non-key activities and omitting key ones can compromise the content validity of these PROMs,^72^ potentially leading to low completion rates, as patients may find irrelevant activities burdensome or important ones missing. Developing outcome measures that prioritize patient needs and eliminate less relevant items could enhance patient-centeredness, potentially reducing the burden on patients and increasing response rates.^73^ Our previous research evaluated the content of outcome measure instruments after hip or knee arthroplasty based on the ICF framework,^71^ and our findings highlight the importance of evaluation of these instruments on key activities identified in this study to determine patient-centeredness of outcome measures.

Comparing our findings with other studies is challenging because there are limited studies on the key functional activities across different time periods following hip or knee arthroplasty. One study explored patient concerns during the initial six weeks following knee arthroplasty,^74^ with concerns overlapping with our study, such as lying down in bed and putting on footwear within two weeks, and going up and down stairs and shopping at six weeks after knee arthroplasty. However, they conducted interviews with only 30 patients. Weiss et al.^75^ examined patient priorities one year after knee arthroplasty, highlighting challenges in functional activities including kneeling, gardening, and squatting for patients. Another study highlighted that more than half of patients expressed dissatisfaction with their ability to perform squatting one year after knee arthroplasty.^76^ Additionally, another study reported that 29.5% of patients continued to experience difficulties with squatting one year after hip arthroplasty, indicating persistent challenges in this activity.^77^ Thus, most activities identified in previous research align with our findings, indicating persistent challenges with these key activities.

### Limitation

Our study had some limitations. While we believe our sample is representative of the hip and knee arthroplasty Canadian population, it is possible that patient priorities could vary across different settings and cultures. In addition, we recruited patients who underwent unilateral primary elective hip or knee arthroplasty for OA, thus our results may not be generalizable to those who underwent arthroplasty for other reasons or bilateral arthroplasty. The recruitment strategy targeted individuals at 2, 6, 13, and 26 weeks following hip or knee arthroplasty, which may restrict the generalizability of the questionnaire to other recovery periods. Nevertheless, as the vast majority of functional improvements typically occur within the first six months post-surgery, ^27,28^ it is unlikely that the results would differ substantially beyond six months. In addition, because participants’ health literacy and educational backgrounds were not documented, the extent to which the findings apply to individuals with lower literacy remains uncertain. That said, our approach where patients were invited to participate rather than self-enroll, along with a high response rate, suggests that the sample likely included individuals across a range of literacy levels. Finally, although cross-sectional studies at different timepoints were conducted rather than a cohort study, we do not believe that this choice impacted our results as there were no statistically significant differences in patient demographics and preoperative function between the four groups.

## Conclusion

This study identified key functional activities that are important to patients, and which they had difficulty performing over the first 26 weeks after hip or knee arthroplasty. The findings indicated that these activities varied over time and were mostly the same for hip and knee arthroplasty surgeries, underscoring the need for tailored postoperative interventions at various stages. These results will help clinicians in selecting the appropriate outcomes measures based on patient needs following hip or knee arthroplasty. Moreover, understanding what matters most to patients enables clinicians to better align interventions with patient needs which might improve outcomes for patients undergoing hip or knee arthroplasty.

## Data Availability

All data produced in the present study are available upon reasonable request to the authors

## Acknowledgments

The authors would like to gratefully acknowledge study participants for their time, patience and contributions. Without their enthusiasm and collaboration, our study would not have been possible.

## References

1. Domínguez-Navarro F, Igual-Camacho C, Silvestre-Muñoz A, Roig-Casasús S, Blasco JM. Effects of balance and proprioceptive training on total hip and knee replacement rehabilitation: a systematic review and meta-analysis. Gait & posture. 2018;62:68–74.

2. Papalia R, Campi S, Vorini F, et al. The role of physical activity and rehabilitation following hip and knee arthroplasty in the elderly. Journal of Clinical Medicine. 2020;9(5):1401.

3. Van Manen MD, Nace J, Mont MA. Management of primary knee osteoarthritis and indications for total knee arthroplasty for general practitioners. Journal of Osteopathic Medicine. 2012;112(11):709–715.

4. van Doormaal MC, Meerhoff GA, Vliet Vlieland TP, Peter WF. A clinical practice guideline for physical therapy in patients with hip or knee osteoarthritis. Musculoskeletal Care. 2020;18(4):575–595.

5. CJRR annual report: Hip and knee replacements in Canada | CIHI. Accessed September 27, 2022. https://www.cihi.ca/en/cjrr-annual-report-hip-and-knee-replacements-in-canada

6. Bandholm T, Wainwright TW, Kehlet H. Rehabilitation strategies for optimisation of functional recovery after major joint replacement. Journal of experimental orthopaedics. 2018;5(1):1–4.

7. Shichman I, Roof M, Askew N, et al. Projections and epidemiology of primary hip and knee arthroplasty in medicare patients to 2040-2060. JBJS Open Access. 2023;8(1):e22.

8. Trujols J, Portella MJ, Iraurgi I, Campins MJ, Siñol N, Cobos JPDL. Patient-reported outcome measures: Are they patient-generated, patient-centred or patient-valued? Journal of Mental Health. 2013;22(6):555–562.

9. Wensing M, Jung HP, Mainz J, Olesen F, Grol R. A systematic review of the literature on patient priorities for general practice care. Part 1: Description of the research domain. Social science & medicine. 1998;47(10):1573–1588.

10. Whitebird RR, Solberg LI, Ziegenfuss JY, et al. Personalized outcomes for hip and knee replacement: the patients point of view. J Patient Rep Outcomes. 2021;5(1):116. doi:10.1186/s41687-021-00393-z

11. Wiering B, De Boer D, Delnoij D. Patient involvement in the development of patient-reported outcome measures: a scoping review. Health Expectations. 2017;20(1):11–23.

12. Wang Y, Yin M, Zhu S, Chen X, Zhou H, Qian W. Patient-reported outcome measures used in patients undergoing total knee arthroplasty: a COSMIN systematic review. Bone & Joint Research. 2021;10(3):203–217.

13. Dowsey MM, Choong PF. The utility of outcome measures in total knee replacement surgery. International journal of rheumatology. 2013;2013.

14. Lützner C, Postler AE, Druschke D, Riedel R, Günther KP, Lange T. Ask patients what they expect! A survey among patients awaiting total hip arthroplasty in Germany. The Journal of Arthroplasty. 2022;37(8):1594–1601.

15. Lange T, Schmitt J, Kopkow C, Rataj E, Günther KP, Lützner J. What do patients expect from total knee arthroplasty? A Delphi consensus study on patient treatment goals. The Journal of arthroplasty. 2017;32(7):2093–2099.

16. Mancuso CA, Salvati EA, Johanson NA, Peterson MG, Charlson ME. Patients’ expectations and satisfaction with total hip arthroplasty. The Journal of arthroplasty. 1997;12(4):387–396. Accessed June 1, 2024.

17. Noble PC, Conditt MA, Cook KF, Mathis KB. The John Insall Award: Patient expectations affect satisfaction with total knee arthroplasty. Clinical Orthopaedics and Related Research (1976-2007). 2006;452:35–43.

18. Schwartz CE, Sprangers MA. Adaptation to Changing Health: Response Shift in Quality-of-Life Research. American Psychological Association; 2000.

19. Razmjou H, Yee A, Ford M, Finkelstein JA. Response shift in outcome assessment in patients undergoing total knee arthroplasty. JBJS. 2006;88(12):2590–2595.

20. Kim TK, Kwon SK, Kang YG, Chang CB, Seong SC. Functional disabilities and satisfaction after total knee arthroplasty in female Asian patients. The Journal of arthroplasty. 2010;25(3):458–464.

21. Von Elm E, Altman DG, Egger M, et al. The Strengthening the Reporting of Observational Studies in Epidemiology (STROBE) Statement: guidelines for reporting observational studies. International journal of surgery. 2014;12(12):1495–1499.

22. Setia MS. Methodology series module 3: Cross-sectional studies. Indian journal of dermatology. 2016;61(3):261.

23. Thompson CB, Panacek EA. Research study designs: Non-experimental. Air medical journal. 2007;26(1):18–22.

24. Karimijashni M, Westby M, Ramsay T, Beaulé PE, Poitras S. Development and content validation of a questionnaire identifying patients’ functional priorities and abilities after hip or knee arthroplasty. Disability and Rehabilitation. 2025;47(8):2146–60.

25. Maloy GC, Kammien AJ, Rubin LE, Grauer JN. Adverse Events After Total Hip Arthroplasty Are Not Sufficiently Characterized by 30 Day Follow-Up: A Database Study. The Journal of Arthroplasty. Published online 2022.

26. Bohl DD, Ondeck NT, Basques BA, Levine BR, Grauer JN. What is the timing of general health adverse events that occur after total joint arthroplasty? Clinical Orthopaedics and Related Research®. 2017;475(12):2952–2959.

27. Wieczorek M, Rotonda C, Guillemin F, Rat AC. What Have We Learned About the Course of Clinical Outcomes After Total Knee or Hip Arthroplasty? Arthritis Care & Research. 2020;72(11):1519–1529.

28. Kennedy DM, Stratford PW, Riddle DL, Hanna SE, Gollish JD. Assessing recovery and establishing prognosis following total knee arthroplasty. Physical therapy. 2008;88(1):22–32.

29. Etikan I, Musa SA, Alkassim RS. Comparison of convenience sampling and purposive sampling. American journal of theoretical and applied statistics. 2016;5(1):1–4.

30. Paulhus DL. Socially desirable responding on self-reports. Encyclopedia of personality and individual differences. 2017;1(5).

31. Burns KE, Duffett M, Kho ME, et al. A guide for the design and conduct of self-administered surveys of clinicians. Cmaj. 2008;179(3):245–252.

32. Kotrlik J, Higgins C. Organizational research: Determining appropriate sample size in survey research appropriate sample size in survey research. Information technology, learning, and performance journal. 2001;19(1):43.

33. Jenny JY, Diesinger Y. The Oxford Knee Score: compared performance before and after knee replacement. Orthopaedics & Traumatology: Surgery & Research. 2012;98(4):409–412.

34. Clement ND, Scott CE, Hamilton DF, MacDonald D, Howie CR. Meaningful values in the forgotten joint score after total knee arthroplasty: minimal clinical important difference, minimal important and detectable changes, and patient-acceptable symptom state. The Bone & Joint Journal. 2021;103(5):846–854.

35. Boonstra AM, Preuper HRS, Balk GA, Stewart RE. Cut-off points for mild, moderate, and severe pain on the visual analogue scale for pain in patients with chronic musculoskeletal pain. Pain®. 2014;155(12):2545–2550.

36. Bjorgul K, Novicoff WM, Saleh KJ. Evaluating comorbidities in total hip and knee arthroplasty: available instruments. Journal of orthopaedics and traumatology. 2010;11(4):203–209.

37. Mendonca D, Warren NA. Perceived and unmet needs of critical care family members. Critical care nursing quarterly. 1998;21(1):58–67.

38. Lei Chang. A Psychometric Evaluation of 4-Point and 6-Point Likert-Type Scales in Relation to Reliability and Validity. Applied Psychological Measurement. 1994;18(3):205–215.

39. Moretti F, van Vliet L, Bensing J, et al. A standardized approach to qualitative content analysis of focus group discussions from different countries. Patient education and counseling. 2011;82(3):420–428.

40. Hsieh HF, Shannon SE. Three Approaches to Qualitative Content Analysis. Qual Health Res. 2005;15(9):1277–1288. doi:10.1177/1049732305276687

41. Lützner C, Postler AE, Druschke D, Riedel R, Günther KP, Lange T. Ask Patients What They Expect! A Survey Among Patients Awaiting Total Hip Arthroplasty in Germany. The Journal of Arthroplasty.

42. Diamond IR, Grant RC, Feldman BM, et al. Defining consensus: a systematic review recommends methodologic criteria for reporting of Delphi studies. Journal of clinical epidemiology. 2014;67(4):401–409.

43. Wylde V, Hewlett S, Learmonth ID, Dieppe P. Persistent pain after joint replacement: prevalence, sensory qualities, and postoperative determinants. PAIN®. 2011;152(3):566–572.

44. Poitras S, Wood KS, Savard J, Dervin GF, Beaule PE. Predicting early clinical function after hip or knee arthroplasty. Bone & Joint Research. 2015;4(9):145–151.

45. Nilsdotter AK, Petersson IF, Roos EM, Lohmander LS. Predictors of patient relevant outcome after total hip replacement for osteoarthritis: a prospective study. Annals of the rheumatic diseases. 2003;62(10):923–930.

46. Nilsdotter AK, Toksvig-Larsen S, Roos EM. Knee arthroplasty: are patients’ expectations fulfilled?: A prospective study of pain and function in 102 patients with 5-year follow-up. Acta Orthopaedica. 2009;80(1):55–61.

47. Stephens MAP, Druley JA, Zautra AJ. Older adults’ recovery from surgery for osteoarthritis of the knee: psychosocial resources and constraints as predictors of outcomes. Health Psychology. 2002;21(4):377.

48. Quintana JM, Escobar A, Arostegui I, et al. Health-related quality of life and appropriateness of knee or hip joint replacement. Archives of internal medicine. 2006;166(2):220–226.

49. Jones CA, Voaklander DC, Johnston DW, Suarez-Almazor ME. Health related quality of life outcomes after total hip and knee arthroplasties in a community based population. The Journal of rheumatology. 2000;27(7):1745–1752.

50. Núñez M, Núñez E, Del Val JL, et al. Health-related quality of life in patients with osteoarthritis after total knee replacement: factors influencing outcomes at 36 months of follow-up. Osteoarthritis and cartilage. 2007;15(9):1001–1007.

51. Oates J, Weston WW, Jordan J. The impact of patient-centered care on outcomes. Fam Pract. 2000;49(9):796–804.

52. Holman H, Lorig K. Patients as partners in managing chronic disease: partnership is a prerequisite for effective and efficient health care. Bmj. 2000;320(7234):526–527.

53. Karimijashni M, Ramsay T, Beaulé PE, Poitras S. Strategies to Manage Poorer Outcomes After Hip or Knee Arthroplasty: A Narrative Review of Current Understanding, Unanswered Questions, and Future Directions. Musculoskeletal Care. 2024;22(3):e1921. doi:10.1002/msc.1921

54. Karimijashni M, Yoo S, Barnes K, et al. Postoperative Rehabilitation Interventions in Patients at Risk of Poorer Outcomes Following Total Knee Arthroplasty: A Systematic Review. Musculoskeletal Care. 2025;23(1):e70054. doi:10.1002/msc.70054

55. Wright JG, Santaguida PL, Young N, Hawker GA, Schemitsch E, Owen JL. Patient preferences before and after total knee arthroplasty. Journal of clinical epidemiology. 2010;63(7):774–782.

56. Salmon P, Hall GM, Peerbhoy D, Shenkin A, Parker C. Recovery from hip and knee arthroplasty: Patients’ perspective on pain, function, quality of life, and well-being up to 6 months postoperatively. Archives of physical medicine and rehabilitation. 2001;82(3):360–366.

57. Gavin JP, Immins T, Wainwright T. Stair negotiation as a rehabilitation intervention for enhancing recovery following total hip and knee replacement surgery. International journal of orthopaedic and trauma nursing. 2017;25:3–10.

58. Kennedy DM, Stratford PW, Robarts S, Gollish JD. Using Outcome Measure Results to Facilitate Clinical Decisions the First Year After Total Hip Arthroplasty. J Orthop Sports Phys Ther. 2011;41(4):232–240.

59. Lenguerrand E, Wylde V, Gooberman-Hill R, et al. Trajectories of pain and function after primary hip and knee arthroplasty: the ADAPT cohort study. PloS one. 2016;11(2):e0149306.

60. Naylor JM, Harmer AR, Heard RC, Harris IA. Patterns of recovery following knee and hip replacement in an Australian cohort. Australian Health Review. 2009;33(1):124–135.

61. Dumenci L, Perera RA, Keefe FJ, et al. Model-based pain and function outcome trajectory types for patients undergoing knee arthroplasty: a secondary analysis from a randomized clinical trial. Osteoarthritis and cartilage. 2019;27(6):878–884.

62. Gandek B, Roos EM, Franklin PD, Ware Jr JE. Item selection for 12-item short forms of the Knee injury and Osteoarthritis Outcome Score (KOOS-12) and Hip disability and Osteoarthritis Outcome Score (HOOS-12). Osteoarthritis and Cartilage. 2019;27(5):746–753.

63. Kennedy DM, Hanna SE, Stratford PW, Wessel J, Gollish JD. Preoperative function and gender predict pattern of functional recovery after hip and knee arthroplasty. The Journal of arthroplasty. 2006;21(4):559–566.

64. Kennedy DM, Stratford PW, Hanna SE, Wessel J, Gollish JD. Modeling early recovery of physical function following hip and knee arthroplasty. BMC Musculoskelet Disord. 2006;7(1):100.

65. Wylde V, Blom AW, Whitehouse SL, Taylor AH, Pattison GT, Bannister GC. Patient-reported outcomes after total hip and knee arthroplasty: comparison of midterm results. The Journal of arthroplasty. 2009;24(2):210–216.

66. Bourne RB, Chesworth B, Davis A, Mahomed N, Charron K. Comparing patient outcomes after THA and TKA: is there a difference? Clinical Orthopaedics and Related Research®. 2010;468(2):542–546.

67. Samuel D, Rowe P, Hood V, Nicol A. The biomechanical functional demand placed on knee and hip muscles of older adults during stair ascent and descent. Gait & posture. 2011;34(2):239–244.

68. Protopapadaki A, Drechsler WI, Cramp MC, Coutts FJ, Scott OM. Hip, knee, ankle kinematics and kinetics during stair ascent and descent in healthy young individuals. Clinical biomechanics. 2007;22(2):203–210.

69. Mizner RL, Snyder-Mackler L. Altered loading during walking and sit-to-stand is affected by quadriceps weakness after total knee arthroplasty. Journal Orthopaedic Research. 2005;23(5):1083–1090.

70. Hyodo K, Masuda T, Aizawa J, Jinno T, Morita S. Hip, knee, and ankle kinematics during activities of daily living: a cross-sectional study. Brazilian journal of physical therapy. 2017;21(3):159–166.

71. Karimijashni M, Abtahi F, Abbasalipour S, et al. Functional Patient-Reported Outcome Measures After Hip or Knee Arthroplasty: A Systematic Review and Content Analysis Using the International Classification of Functioning, Disability, and Health. Arthritis Care & Research. 2024;76(12):1703–1722. doi:10.1002/acr.25413

72. Terwee CB, Prinsen CAC, Chiarotto A, et al. COSMIN methodology for evaluating the content validity of patient-reported outcome measures: a Delphi study. Qual Life Res. 2018;27(5):1159–1170.

73. Rolstad S, Adler J, Rydén A. Response burden and questionnaire length: is shorter better? A review and meta-analysis. Value in Health. 2011;14(8):1101–1108.

74. Rastogi R, Davis AM, Chesworth BM. A cross-sectional look at patient concerns in the first six weeks following primary total knee arthroplasty. Health Qual Life Outcomes. 2007;5(1):48.

75. Weiss JM, Noble PC, Conditt MA, et al. What functional activities are important to patients with knee replacements? Clinical Orthopaedics and Related Research®. 2002;404:172–188.

76. Du H, Tang H, Gu JM, Zhou YX. Patient satisfaction after posterior-stabilized total knee arthroplasty: a functional specific analysis. The Knee. 2014;21(4):866–870.

77. Harada S, Hamai S, Gondo H, Higaki H, Ikebe S, Nakashima Y. Squatting after total hip arthroplasty: patient-reported outcomes and in vivo three-dimensional kinematic study. The Journal of Arthroplasty. 2022;37(4):734–741.

