## Appendix for "Which functional activities matter most to the patients during the first six months after hip or knee arthroplasty: a survey study"

**Appendix A**. Demographic characteristics of patients who underwent hip arthroplasty at different time points.

| | | **Within 2 weeks postop (N=127)** | **6 weeks postop (N=124)** | **13 weeks postop (N=129)** | **26 weeks postop (N=123)** | **P-value** |
|---|---|---|---|---|---|---|
| **Surgery type** | Total hip arthroplasty | 106 | 109 | 117 | 114 | - |
| | Hip resurfacing | 21 | 15 | 12 | 9 | - |
| **Age** Mean (SD) (range) | | 63.71 (9.82) (41-86) | 65.15 (10.35) (40-87) | 66.09 (10.2) (41-88) | 65.14 (9.8) (40-86) | 0.26 |
| **Sex** (male, female) | | (64, 63) | (66,58) | (57,72) | (58,66) | 0.5 |
| **Gender (**man, woman) | | (64, 63) | (66,58) | (57,72) | (58,66) | 0.5 |
| **BMI** Mean (SD) (range) | | 28.56 (5.6) (17.3-53.16) | 28.16 (5.5) (19.4-48.7) | 27.44 (5.7) (16 -55.3) | 28.92 (5.39) (19.2-45.9) | 0.57 |
| **Preop OHS** Mean (SD) | | 21.84 (8.9) | 23.16 (9.01) | 21.83 (8.9) | 21 (9.65) | 0.3 |
| **Ethnicity** | White | 116 | 113 | 119 | 115 | 0.41 |
| | First Nation, Métis,Inuk(Inuit) | 1 | 0 | 0 | 1 | |
| | South Asian | 0 | 1 | 1 | 0 | |
| | Chinese | 3 | 2 | 0 | 1 | |
| | Black | 1 | 1 | 1 | 1 | |
| | Latin American | 2 | 0 | 0 | 0 | |
| | Arab | 1 | 2 | 1 | 0 | |
| | Japanese | 0 | 0 | 0 | 1 | |
| | Prefer not to say | 3 | 5 | 4 | 4 | |
| **ASA score** | ASA=1 | 7 | 1 | 12 | 8 | 0.043 |
| | ASA=2 | 51 | 47 | 38 | 52 | |
| | ASA=3 | 64 | 66 | 67 | 58 | |
| | ASA=4 | 4 | 9 | 12 | 5 | |
| **Receiving precautions** (Yes, No) | | (118, 9) | (77, 43) | (35, 93) | (27, 95) | **<0.001** |
| **Pain** Mean (SD) | | 39.74 (22.1) | 18.8 (20) | 14.91(17.9) | 14.57 (20.7) | **<0.001** |
| **Problems in other joints (yes, no)** | | (78, 46) | (77, 44) | (92, 33) | (86, 34) | 0.23 |
| **Problems in other joints** | Right Hip (yes) | 23 (18.1%) | 23 (18.5%) | 21 (16.3%) | 18 (14.6%) | 0.83 |
| | Left hip (yes) | 29 (22.8%) | 17(13.7%) | 26 (20.2%) | 14 (11.4%) | 0.05 |
| | Right knee (yes) | 32 (25.2%) | 26 (21%) | 30 (23.2%) | 31 (25.2%) | 0.84 |
| | Left knee (yes) | 25 (19.7%) | 24 (19.4%) | 28 (21.7%) | 25 (20.3%) | 0.96 |
| | Right foot (yes) | 8 (6.3 %) | 5 (4%) | 5 (5.4%) | 12 (9.7%) | 0.3 |
| | Left foot (yes) | 5 (3.9%) | 5 (4%) | 4 (3.1%) | 13 (10.6%) | 0.03 |
| | Low back (yes) | 24 (18.9%) | 30 (24.2%) | 47 (36.4%) | 44 (35.7%) | 0.003 |
| | Neck (yes) | 8 (6.3%) | 7 (5.6%) | 14 (10.9%) | 17 (13.8%) | 0.08 |
| | Shoulder (yes) | 14 (11%) | 16 (12.9%) | 22 (17.1%) | 33(26.8%) | **0.004** |
| **Walking aids** | Wheelchair (yes) | 3 (2.4%) | 1 (0.8%) | 1(0.8%) | 0 | 0.29 |
| | Walker (yes) | 73(57.5%) | 23 (18.5%) | 14(10.9%) | 12 (9.75%) | **<0.001** |
| | Cane (yes) | 58 (45.7%) | 55 (44.4%) | 39(30.2%) | 19 (15.4%) | **<0.001** |
| | Crutches (yes) | 47 (37%) | 12 (9.7%) | 1 (0.8%) | 1 (0.8%) | **<0.001** |
| | Scooter (yes) | 1 (0.8%) | 2 (1.6%) | 1(0.8%) | 0 | 0.56 |
| | Other walking aids (yes) | 2 (1.6%) | 5 (4%) | 2 (1.6%) | 4 (3.2%) | 0.51 |
| | No using aids (yes) | 10 (7.9%) | 52 (41.9%) | 86 (66.7%) | 97 (78.9%) | **<0.001** |
| **Living with whom** alone, family | | 17, 110 | 26, 97 | 25, 104 | 29, 94 | 0.21 |

| | | | | | | |
|---|---|---|---|---|---|---|
| **Having family** yes, no | | 108,18 | 97,23 | 101,27 | 97, 24 | 0.53 |
| **Dependency of others** yes, no | | 19, 107 | 17,104 | 23,105 | 30,93 | 0.2 |
| **Place of residence** | House | 105 | 105 | 110 | 101 | 0.84 |
| | Apartment | 18 | 16 | 17 | 20 | |
| | Retirement home | 2 | 0 | 1 | 0 | |
| | Other | 1 | 2 | 1 | 2 | |
| **Level of activity** | Sedentary | 32 | 21 | 21 | 22 | 0.06 |
| | Light physical | 17 | 19 | 19 | 31 | |
| | Heavy physical | 14 | 10 | 6 | 7 | |
| | Retired | 64 | 73 | 81 | 63 | |

Some values may contain slightly missing data

**Appendix B**. Demographic characteristics of patients who underwent knee arthroplasty at different time points.

| | | Within 2 weeks postop (N=112) | 6 weeks postop (N=112) | 13 weeks postop (N=116) | 26 weeks postop (N=113) | P-value |
|---|---|---|---|---|---|---|
| **Surgery Type** | Total knee arthroplasty | 111 | 107 | 107 | 108 | - |
| | Unicompartmental knee arthroplasty | 1 | 5 | 9 | 5 | - |
| **Age** Mean (SD) (range) | | 67.2 (8.86) (44-87) | 68.35 (8.0) (49-87) | 69.03(8.8) (44-90) | 68.32 (8.9) (48-88) | 0.37 |
| **Sex** (male, female) | | (57, 55) | (53,62) | (56,59) | (55,58) | 0.79 |
| **Gender** (man, woman) | | (57, 55) | (53,62) | (56,59) | (55,58) | 0.79 |
| **BMI** Mean (SD) (range) | | 32.8 (6) (17.5-50.22) | 31.21 (6.4) (21.60-51.2) | 30.33 (6.2) (19.71-54.38) | 31.1 (7) (20.53-54.9) | 0.39 |
| **Preop OKS** Mean (SD) | | 23.5 (8.5) | 22.8 (7.1) | 24.7 (7.7) | 22.9 (7.9) | 0.61 |
| **Ethnicity** | White | 98 | 95 | 103 | 105 | 0.32 |
| | First Nation, Métis, Inuk(Inuit) | 2 | 1 | 0 | 0 | |
| | South Asian | 2 | 5 | 2 | 1 | |
| | Chinese | 1 | 0 | 0 | 1 | |
| | Black | 1 | 1 | 0 | 2 | |
| | Filipino | 1 | 1 | 0 | 0 | |
| | Arab | 3 | 0 | 2 | 0 | |
| | Prefer not to say | 4 | 7 | 7 | 3 | |
| **ASA score** | ASA=1 | 1 | 1 | 4 | 3 | 0.61 |
| | ASA=2 | 42 | 34 | 32 | 32 | |
| | ASA=3 | 65 | 75 | 75 | 73 | |
| | ASA=4 | 2 | 5 | 4 | 5 | |
| **Pain** Mean (SD) | | 52.15 (22.05) (0-98) | 33.14 (24.24) (0-95) | 27.7 (22.8) (0-81) | 22.7 (21.7) (0-85) | **<0.001** |
| **Problems in other joints (yes, no)** | | (44, 64) | (76, 33) | (79, 32) | (85, 27) | 0.06 |
| **Problems in other joints n** | Right Hip (yes) | 16 (14.5%) | 18 (15.7%) | 16 (13.9%) | 16 (14.2%) | 0.96 |
| | Left hip (yes) | 10 (9.1%) | 16 (13.9%) | 16 (13.9%) | 14 (12.4%) | 0.73 |
| | Right knee (yes) | 29 (26.4%) | 32 (27.8%) | 30 (26.1%) | 30 (26.5%) | 0.99 |
| | Left knee (yes) | 31 (28.2%) | 29 (25.2%) | 31 (27.0%) | 35 (31%) | 0.8 |
| | Right foot (yes) | 8 (7.3%) | 14 (12.2%) | 11 (9.6 %) | 8 (7.1%) | 0.45 |
| | Left foot (yes) | 6 (5.5%) | 7 (6.1%) | 10 (8.7%) | 8 (7.1%) | 0.81 |

| | | | | | | |
|---|---|---|---|---|---|---|
| | Low back (yes) | 17 (15.5%) | 27 (23.5%) | 26 (22.6%) | 30 (26.5%) | 0.24 |
| | Neck (yes) | 4 (3.6%) | 10 (8.7%) | 10 (8.7%) | 16 (14.2%) | 0.05 |
| | Shoulder (yes) | 7 (6.4%) | 16 (13.9%) | 21 (18.3%) | 22 (19.5%) | **0.03** |
| **Walking aids** | | | | | | |
| **Walking aids** | Wheelchair (yes) | 3 (2.7%) | 0 | 0 | 1 (0.9%) | 0.1 |
| | Walker (yes) | 63 (57.3%) | 14 (12.2%) | 11 (9.6%) | 6 (5.3%) | **<0.001** |
| | Cane (yes) | 64 (58.2%) | 49 (42.6%) | 26 (22.6%) | 18(15.9%) | **<0.001** |
| | Crutches (yes) | 26 (23.6%) | 4 (3.5%) | 0 | 0 | **<0.001** |
| | Scooter (yes) | 0 | 0 | 0 | 0 | - |
| | Other walking aids (yes) | 2 (1.8%) | 4 (3.5%) | 4 (3.5%) | 5 (4.4%) | 0.75 |
| | No using aids | 8 (7.3%) | 57 (49.6%) | 82 (71.3 %) | 86(77.9%) | **<0.001** |
| **Living with whom** alone, family | | 21,88 | 28, 87 | 27,88 | 25,88 | .9 |
| **Having family** yes, no | | 94, 17 | 95, 19 | 98, 17 | 89, 23 | 0.65 |
| **Dependency of others** yes, no | | 10, 98 | 18, 93 | 18, 95 | 18,95 | 0.42 |
| **Place of residence** | House | 95 | 95 | 98 | 95 | 0.64 |
| | Apartment | 15 | 19 | 16 | 15 | |
| | Retirement home | 0 | 0 | 1 | 2 | |
| | Other | 0 | 0 | 0 | 1 | |
| **Level of activity** | Sedentary | 12 | 14 | 17 | 13 | 0.16 |
| | Light physical | 19 | 14 | 10 | 23 | |
| | Heavy physical | 13 | 8 | 5 | 8 | |
| | Retired | 66 | 75 | 82 | 67 | |

Some values may contain slightly missing data

**Appendix C**. Demographic characteristics of patients who underwent hip or knee arthroplasty.

| | | **Knee arthroplasty (450)** | **Hip arthroplasty (503)** | **P-value** |
|---|---|---|---|---|
| **Age** (Mean (SD), (range) | | 68.21 (8.6) (44-90) | 65 (10.07) (40-88) | **<0.001** |
| **Sex** (male, female) | | (219,231) | (246, 257) | 0.9 |
| **Gender** (man, woman) | | (219,231) | (246, 257) | 0.9 |
| **BMI** (Mean (SD), Median (range) | | 31.09 (6.4) (17.5-54.9) | 28.26(5.6) (16 -55.3) | **<0.001** |
| **OKS or OHS** | | 24.3 (7.8) | 21.9 (9.1) | 0.015 |
| **Ethnicity** | White | 398 | 463 | 0.09 |
| | First Nation, Métis, Inuk(Inuit) | 3 | 2 | |
| | South Asian | 10 | 1 | |
| | Chinese | 2 | 6 | |
| | Black | 4 | 4 | |
| | Filipino | 2 | 0 | |
| | Lati American | 1 | 2 | |
| | Arab | 5 | 4 | |
| | Japanese | 0 | 1 | |
| | Prefer not to say | 25 | 19 | |
| **ASA score** | ASA=1 | 9 | 28 | **<0.001** |
| | ASA=2 | 140 | 188 | |
| | ASA=3 | 285 | 255 | |

| | | | | |
|---|---|---|---|---|
| | ASA=4 | 16 | 30 | |
| **Receiving Precautions** (Yes, %) | | - | 257 (51.1%) | - |
| **Pain** (Mean (SD) | | 33.5 (25.14) | 21.9 (22.7) | **<0.001** |
| **Problems in other joints** (Yes, No) | | (303, 136) | (333, 157) | 0.60 |
| **Problems in other joints** | Right Hip (yes) | 66 (14.7%) | 85 (16.9%) | 0.35 |
| | Left hip (yes) | 54 (12%) | 86 (17.1%) | **0.03** |
| | Right knee (yes) | 121 (26.9%) | 119 (23.7%) | 0.25 |
| | Left knee (yes) | 125 (27.8%) | 102 (20.3%) | **0.007** |
| | Right foot (yes) | 41 (9.1%) | 32 (6.4%) | 0.11 |
| | Left foot (yes) | 31 (6.9%) | 27 (5.4 %) | 0.33 |
| | Low back (yes) | 99 (22%) | 145 (28.8%) | 0.02 |
| | Neck (yes) | 40 (8.9%) | 46 (9.1%) | 0.9 |
| | Shoulder (yes) | 66 (14.7%) | 85 (16.9%) | 0.35 |
| **Walking aids** | Wheelchair (yes) | 4 (.9%) | 4 (1%) | 0.87 |
| | Walker (yes) | 93 (20.7%) | 122 (24.3%) | 0.2 |
| | Cane (yes) | 155 (34.4%) | 171 (32.3%) | 0.88 |
| | Crutches (yes) | 30 (6.7%) | 61(12.1%) | **0.004** |
| | Scooter (yes) | 0 | 4 (.8%) | 0.06 |
| | Other walking aids (yes) | 15 (3.3%) | 13 (2.6%) | 0.49 |
| | No using aids | 233 (51.8%) | 245 (48.7%) | 0.34 |
| **Living with whom** (alone, family) | | (98,351) | (97,405) | 0.34 |
| **Having family** (yes,no) | | (372,75) | (403,92) | 0.47 |
| **Dependency of others** (yes,no) | | (64,381) | (89,409) | 0.23 |
| **Place of residence** | House | 379 | 421 | 0.52 |
| | Apartment | 64 | 71 | |
| | Retirement home | 3 | 3 | |
| | Other | 1 | 5 | |
| **Level of activity** | Sedentary, n (%) | 56 (12.4%) | 96 (19.1%) | **0.02** |
| | Light physical, n (%) | 66 (14.7%) | 86 (17.1%) | |
| | Heavy physical, n (%) | 34 (7.6%) | 37 (7.4%) | |
| | Retired, n (%) | 285 (63.3%) | 281 (55.9%) | |

**Appendix D.** Importance and difficulty of key activities for hip and knee arthroplasty **within two weeks.**

| | **Within 2 weeks postoperative** | | | |
|---|---|---|---|---|
| **Activity** | **Importance%_Hip** | **Importance%_Knee** | **Difficulty%_Hip** | **Difficulty%_Knee** |
| **Changing and holding a body position** | | | | |
| Lying down in bed and getting up | **97.7** | **94.4** | **11.8** | **15.7** |
| Rolling over | **81.9** | **84.3** | **25.2** | **32.4** |
| Hold a lying position during the day | 63.8 | 60.2 | 17.3 | 24.1 |
| Squatting down | 63 | 51.8 | 44.1* | 69.4 |
| Hold a squatting position | 44.1 | 42.6 | 52.7* | 76.9 |
| Kneeling down on the ground | 33 | 35.2 | 53.6* | 93.5 |
| Hold a kneeling position | 30.7 | 27.8 | 50.4* | 92.6 |
| Sitting on a chair | 92.2 | 95.4 | 3.2 | 9.3 |
| Hold a sitting position | **88.2** | **91.6** | **15.7*** | **37.9** |
| Rising from a chair | **92.1** | **89.8** | **9.4*** | **19.5** |
| Hold a standing position | **85** | **80.6** | **33.1*** | **44.5** |
| Bending over or to the side | 63 | 71.3 | 35.5* | 37.1 |
| Shifting the body from side to side | 63 | 66.6 | 18.9 | 25.9 |
| **Lifting and carrying objects** | | | | |
| Lifting a light object from the floor | 62.2 | 63.9 | 33.9* | 32.4 |
| Lifting a heavy object from the floor | 29.1 | 39.8 | 51.9* | 70.4 |
| Putting down objects on the floor | 40.9 | 49 | 36.2* | 45.4 |
| Carrying in the hands | 57.5 | 60.2 | 31.5 | 38.9 |
| Carrying in the arms | 47.2 | 51.9 | 41.7 | 50 |
| Carrying on shoulders or back | 23.6 | 25 | 18.1 | 20.3 |
| **Walking** | | | | |
| Walking short distances | **80.3** | **75.9** | **20.5*** | **39.8** |
| Walking long distances | 39.4 | 39.8 | 50.4* | 69.4 |
| Walking on different surfaces | 48.8 | 44.5 | 40.2* | 60.2 |
| Walking around obstacles | 45.7 | 44.4 | 27.6 | 41.7 |
| **Moving around** | | | | |
| Going up stairs | **76.4** | **72.2** | **26.8*** | **45.4** |
| Going down stairs | **77.2** | **72.3** | **26*** | **49.1** |
| Going up a curb or step | 70.8 | 72.2 | 14.9* | 28.7 |
| Jumping | 15 | 14.9 | 52.7* | 76.9 |
| **Moving around using transportation** | | | | |
| Getting in and out of a car | 69.3 | 72.2 | 29.1* | 40.8 |
| Using public transportation | 10.2 | 11.1 | 14.1* | 35.2 |
| Biking | 15.8 | 20.4 | 37* | 56.5 |
| Driving vehicle | 49.6 | 55.5 | 40.1 | 57.4 |
| **Self-care** | | | | |
| Washing lower body parts | **89** | **88.9** | **57.5*** | **39.8** |
| Washing whole body | **94.5** | **91.6** | **29.2** | **23.2** |
| Drying your body | **93.7** | **90.7** | **24.4** | **18.6** |
| Going to the toilet | 98.4 | 95.4 | 11 | 14.8 |
| Putting on clothes | **93.7** | **90.** | **26** | **19.4** |
| Taking off clothes | 95.3 | 90.8 | 14.2 | 14.8 |
| Putting on footwear | **82.6** | **84.3** | **57.5** | **42.6** |
| Taking off footwear | **84.3** | **86.1** | **39.4** | **27.7** |
| **Domestic life and Intimate relationships** | | | | |
| Going shopping | 31.5 | 31.4 | 38.6 | 57.4 |

| | | | | |
|---|---|---|---|---|
| Doing housework | 50.4 | 46.3 | 52 | 57.4 |
| Assisting others | 26 | 25.9 | 35.4 | 44.4 |
| Sexual relationships | 38.6 | 33.4 | 48 | 50.9 |
| **Employment** | | | | |
| Paid job | 29.1 | 27.7 | 32.3 | 36.1 |
| Unpaid work | 18.9 | 26.8 | 38.6 | 46.3 |
| **Community, social and civic life** | | | | |
| Socializing /Participating in community life | 54.3 | 59.2 | 44.9 | 58.4 |
| Sports | 42.6 | 37.9 | 70.1 | 81.5 |
| Running/jogging | 15.7 | 5.6 | 68.5* | 75 |
| Hobbies/play/Crafts/Arts/culture | 59.8 | 45.4 | 30.7 | 43.5 |

* Difference between hip and knee is significant (p<0.05). Bold numbers indicate key activities.

**Appendix E.** Importance and difficulty of key activities for hip and knee arthroplasty **at six weeks**.

| | 6 weeks postoperative | | | |
|---|---|---|---|---|
| **Activity** | **Importance%_Hip** | **Importance%_Knee** | **Difficulty%_Hip** | **Difficulty%_Knee** |
| **Changing and holding a body position** | | | | |
| Lying down in bed and getting up | 95.2 | 96.5 | 2.4 | 2.6 |
| Rolling over | 88.7 | 91.3 | 11.3 | 7.8 |
| Hold a lying position during the day | 49.2 | 55.7 | 15.3 | 20 |
| Squatting down | **79** | **73.1** | **18.5*** | **32.2** |
| Hold a squatting position | 56.5 | 54.8 | 37.9 | 54.8 |
| Kneeling down on the ground | 55.7 | 50.4 | 38.7 * | 71.3 |
| Hold a kneeling position | 43.5 | 46.9 | 36.3* | 76.6 |
| Sitting on a chair | 98.4 | 100 | .8 | 2.6 |
| Hold a sitting position | **96.8** | **95.6** | **10.5** | **20.9** |
| Rising from a chair | 97.6 | 98.3 | 4.8 | 7 |
| Hold a standing position | 93.5 | 96.5 | 11.3* | 14.8 |
| Bending over or to the side | 91.9 | 95.7 | 14.9* | 7.8 |
| Shifting the body from side to side | 75.8 | 77.4 | 4.8 | 5.2 |
| **Lifting and carrying objects** | | | | |
| Lifting a light object from the floor | 82.2* | 93.9 | 12.9* | 3.5 |
| Lifting a heavy object from the floor | 62.1 | 67.8 | 35.4 | 31.3 |
| Putting down objects on the floor | **75** | **79.1** | **19.4** | **7.8** |
| Carrying in the hands | 88.7 | 91.3 | 12.1 | 11.3 |
| Carrying in the arms | **80.6** | **84.3** | **17.8** | **13** |
| Carrying on shoulders or back | 54.8 | 46.1 | 6.4 | 4.3 |
| **Walking** | | | | |
| Walking short distances | 95.1 | 94.8 | 13.7 | 6.1 |
| Walking long distances | 70.9 | 70.4 | 33.1 | 28.7 |
| Walking on different surfaces | **79** | **80** | **24.2** | **26.1** |
| Walking around obstacles | 78.2 | 77.4 | 10.5 | 8.7 |
| **Moving around** | | | | |
| Going up stairs | **91.1** | **87.8** | **16.7*** | **19.2** |
| Going down stairs | **95.8** | 85.2 | 12.1* | 26.9 |
| Going up a curb or step | 90.4 | 93.9 | 4.8 | 8.7 |

| | | | | |
|---|---|---|---|---|
| Jumping | 21 | 24.3 | 29.8 | 40.9 |
| **Moving around using transportation** | | | | |
| Getting in and out of a car | 94.3 | 95.7 | 4.8 | 7.8 |
| Using public transportation | 22.6 | 13.9 | 4.8 | 6.1 |
| Biking | 27.4 | 35.6 | 18.5* | 27 |
| Driving vehicle | 87.1 | 92.2 | 8.1 | 11.3 |
| **Self-care** | | | | |
| Washing lower body parts | **92.8** | 96.6 | **21.7*** | 9.6 |
| Washing whole body | 96.7 | 100 | 4 | 7.9 |
| Drying your body | 96 | 96.5 | 4.8 | 3.5 |
| Going to the toilet | 100 | 97.4 | 1.6 | 4.3 |
| Putting on clothes | 99.1 | 98.3 | 4 | 3.5 |
| Taking off clothes | 98.4 | 97.4 | 3.2 | 3.5 |
| Putting on footwear | **99.2** | **97.4** | **25** | **13** |
| Taking off footwear | **98.4** | **96.5** | **17.7** | **8.7** |
| **Domestic life and Intimate relationships** | | | | |
| Going shopping | **76.6** | **76.5** | **10.5** | **18.3** |
| Doing housework | **87.1** | **86.9** | **20.1** | **20.9** |
| Assisting others | 43.5 | 51.3 | 8.8 | 19.1 |
| Sexual relationships | 52.4 | 37.9 | 14.5 | 19.1 |
| **Employment** | | | | |
| Paid job | 35.5 | 28.7 | 14.5 | 14.7 |
| Unpaid work | 27.4 | 33.9 | 12.9 | 16.5 |
| **Community, social and civic life** | | | | |
| Socializing/Participating in community life | **77.5** | **77.4** | **7.2** | **16.5** |
| Sports | 58.9 | 55.6 | 51.6 | 47.9 |
| Running/jogging | 17.8 | 18.2 | 39.6* | 45.2 |
| Hobbies/play/Crafts/Arts/culture | 69.3 | 75.6 | 9.7 | 10.5 |

* Difference between hip and knee is significant (p<0.05). Bold numbers indicate key activities.

**Appendix F.** Importance and difficulty of key activities for hip and knee arthroplasty **at 13 weeks.**

| | **13 weeks postoperative** | | | |
|---|---|---|---|---|
| **Activity** | **Importance%_Hip** | **Importance%_Knee** | **Difficulty%_Hip** | **Difficulty%_Knee** |
| **Changing and holding a body position** | | | | |
| Lying down in bed and getting up | 96.2 | 93.1 | 0 | 1.7 |
| Rolling over | 93 | 87.9 | 3.1 | 1.7 |
| Hold a lying position during the day | 54.3 | 53.1 | 13.2 | 18.3 |
| Squatting down | **88.4*** | **79.1** | **14** | **20.9** |
| Hold a squatting position | 68.2 | 62.6 | 25.6 | 33.9 |
| Kneeling down on the ground | 72.8* | 56.5 | 34.9* | 62.7 |
| Hold a kneeling position | 62.8 | 48.7 | 23.3* | 62.6 |
| Sitting on a chair | 96.1 | 91.3 | .8 | 0 |
| Hold a sitting position | **91.5** | **92.1** | **8.5** | **15.6** |
| Rising from a chair | 94.6 | 92.2 | 6.2 | 3.5 |
| Hold a standing position | 94.6 | 90.4 | 7.8 | 13.9 |
| Bending over or to the side | 88.4 | 85.2 | 9.4 | 7 |
| Shifting the body from side to side | 75.2 | 76.6 | 2.3 | 1.7 |

| | | | | |
|---|---|---|---|---|
| **Lifting and carrying objects** | | | | |
| Lifting a light object from the floor | 90.7 | 86.9 | 6.3 | 2.6 |
| Lifting a heavy object from the floor | **76.8** | **74.8** | **27.1** | **19.1** |
| Putting down objects on the floor | 86.9 | 80.9 | 8.6 | 5.2 |
| Carrying in the hands | 93 | 88.7 | 3.1 | 3.5 |
| Carrying in the arms | 83.7 | 82.6 | 7.8 | 9.5 |
| Carrying on shoulders or back | 56.6 | 47.8 | 6.2 | 3.5 |
| **Walking** | | | | |
| Walking short distances | 96.1 | 94.8 | 1.6 | 1.7 |
| Walking long distances | **77.5** | **79.2** | **20.2** | **22.6** |
| Walking on different surfaces | 86.1 | 81.7 | 14* | 13 |
| Walking around obstacles | 83.7 | 80.9 | 6.3 | 5.2 |
| **Moving around** | | | | |
| Going up stairs | 94.6 | 91.3 | 9.3 | 11.3 |
| Going down stairs | 93 | **90.5** | 5.5* | **19.1** |
| Going up a curb or step | 94.5 | 89.6 | 4.7 | 3.5 |
| Jumping | 31 | 22.6 | 23.3* | 37.3 |
| **Moving around using transportation** | | | | |
| Getting in and out of a car | 97.6 | 93 | 3.9 | 5.3 |
| Using public transportation | 26.4 | 27.8 | 3.2 | 5.2 |
| Biking | 41.1 | 46.1 | 8.5 | 17.4 |
| Driving vehicle | 89.9 | 89.6 | 5.5* | 0.9 |
| **Self-care** | | | | |
| Washing lower body parts | 97.7 | 96.4 | 11.7 | 5.2 |
| Washing whole body | 99.3 | 93.9 | 3.1 | 0.9 |
| Drying your body | 96.9 | 93 | 1.6* | 0 |
| Going to the toilet | 99.3 | 95.6 | 2.3 | 0 |
| Putting on clothes | 98.5 | 96.6 | 1.6 | 0 |
| Taking off clothes | 99.2 | 94.8 | 0.8 | 0 |
| Putting on footwear | 97.7 | 95.6 | 14.7 | 7 |
| Taking off footwear | 96.1 | 94.8 | 9.4 | 6.1 |
| **Domestic life and Intimate relationships** | | | | |
| Going shopping | 80.6 | 80.8 | 6.3 | 3.5 |
| Doing housework | 90 | 86.9 | 14* | 5.2 |
| Assisting others | 55.8 | 59.1 | 6.2 | 3.5 |
| Sexual relationships | 53.5 | 45.2 | 10.2 | 11.3 |
| **Employment** | | | | |
| Paid job | 38 | 27.8 | 9.4 | 7.8 |
| Unpaid work | 44.9 | 41.7 | 10.1 | 5.2 |
| **Community, social and civic life** | | | | |
| Socializing/Participating in community life | 87.6 | 81.8 | 6.2 | 2.6 |
| Sports | 66.7 | 56.5 | 22.5 | 29.6 |
| Running/jogging | 20.9 | 20.9 | 31 | 36.5 |
| Hobbies/play/Crafts/Arts/culture | 74.4 | 77.4 | 9.3 | 7.9 |

* Difference between hip and knee is significant ($p<0.05$). Bold numbers indicate key activities.

**Appendix G.** Importance and difficulty of key activities for hip and knee arthroplasty **at 26 weeks.**

| | **26 weeks postoperative** | | | |
|---|---|---|---|---|
| | **Importance%_Hip** | **Importance%_Knee** | **Difficulty%_Hip** | **Difficulty%_Knee** |
| **Changing and holding a body position** | | | | |
| Lying down in bed and getting up | 95.2 | 92.1 | 3.2* | 0 |
| Rolling over | 88.7 | 90.3 | 4.8 | 4.4 |
| Hold a lying position during the day | 51.6 | 55.8 | 10.5 | 13.3 |
| Squatting down | **84.7** | **81.5** | **17.7** | **23** |
| Hold a squatting position | 69.3 | 69 | 26.6 | 29.2 |
| Kneeling down on the ground | 69.4 | 65.5 | 25* | 71.7 |
| Hold a kneeling position | 56.4 | 57.5 | 21* | 64.6 |
| Sitting on a chair | 96 | 92.9 | 2.4 | 0 |
| Hold a sitting position | 93.5 | 92.1 | 9.7 | 7.1 |
| Rising from a chair | 95.9 | 91.1 | 5.6 | 5.3 |
| Hold a standing position | 96 | 90.3 | 9.7 | 8.9 |
| Bending over or to the side | 91.9 | 90.3 | 10.5 | 10.6 |
| Shifting the body from side to side | 78.2 | 74.3 | 4.8 | 2.7 |
| **Lifting and carrying objects** | | | | |
| Lifting a light object from the floor | 94.3 | 90.2 | 4 | 6.2 |
| Lifting a heavy object from the floor | **81.4** | **80.6** | **16.9** | **18.6** |
| Putting down objects on the floor | 87.9 | 84.1 | 4 | 8.8 |
| Carrying in the hands | 93.5 | 93.8 | 5.6 | 7.1 |
| Carrying in the arms | 92 | 88.5 | 7.2 | 8.9 |
| Carrying on shoulders or back | 66.1 | 58.4 | 2.4 | 5.3 |
| **Walking** | | | | |
| Walking short distances | 95.2 | 95.6 | 4 | 4.5 |
| Walking long distances | **81.4** | **80.5** | **14.5** | **17.7** |
| Walking on different surfaces | **88.7** | **86.8** | **13.7** | **20.4** |
| Walking around obstacles | 88.7 | 84 | 4.8 | 8 |
| **Moving around** | | | | |
| Going up stairs | **97.6** | **92.9** | **8.9*** | **15** |
| Going down stairs | **96.8** | **94.7** | **5.6*** | **19.5** |
| Going up a curb or step | 95.2 | 91.2 | 4.8 | 9.7 |
| Jumping | 33.1 | 23 | 24.2* | 40.7 |
| **Moving around using transportation** | | | | |
| Getting in and out of a car | 97.6 | 95.5 | 4.8 | 8.9 |
| Using public transportation | 28.2 | 28.3 | 2.4 | 4.5 |
| Biking | 45.9 | 39 | 9.7 | 16.8 |
| Driving vehicle | 92.8 | 92.1 | 2.4 | 4.4 |
| **Self-care** | | | | |
| Washing lower body parts | 96.8 | 93.8 | 5.6 | 3.6 |
| Washing whole body | 95.9 | 96.5 | 3.2 | 3.5 |
| Drying your body | 97.6 | 95.6 | 2.4 | 0.9 |
| Going to the toilet | 100 | 96.4 | 0.8* | 2.7 |
| Putting on clothes | 100 | 96.5 | 5.6 | 3.5 |
| Taking off clothes | 100 | 96.4 | 3.2 | 0.9 |
| Putting on footwear | 97.6 | 95.6 | 8.9 | 11.5 |
| Taking off footwear | 97.6 | 93.8 | 6.4 | 8 |
| **Domestic life and Intimate relationships** | | | | |
| Going shopping | 90.4 | 87.6 | 4.8 | 3.6 |
| Doing housework | 94.4 | 94.7 | 6.4 | 8 |

| | | | | |
|---|---|---|---|---|
| Assisting others | 61.3 | 63.8 | 4.8 | 7.1 |
| Sexual relationships | 62.9 | 52.2 | 11.3 | 12.4 |
| **Employment** | | | | |
| Paid job | 51.6 | 40.7 | 8.1 | 4.5 |
| Unpaid work | 53.2 | 37.2 | 6.4 | 4.5 |
| **Community, social and civic life** | | | | |
| Socializing/Participating in community life | 89.5 | 85.9 | 5.6 | 4.5 |
| Sports | 70.2 | 69.9 | 24.2 | 30.1 |
| Running/jogging | 21 | 21.2 | 35.5 | 49.5 |
| Hobbies/play/Crafts/Arts/culture | 84.7 | 72.5 | 5.6* | 9.8 |

* Difference between hip and knee is significant ($p<0.05$). Bold numbers indicate key activities.
